# Implementation of medical therapy in different stages of heart failure with reduced ejection fraction

**DOI:** 10.1101/2025.04.21.25326175

**Authors:** Noel G Panagiotides, Annika Weidenhammer, Suriya Prausmüller, Marc Stadler, Georg Spinka, Gregor Heitzinger, Henrike Arfsten, Guido Strunk, Philipp E Bartko, Georg Goliasch, Christian Hengstenberg, Martin Hülsmann, Noemi Pavo

## Abstract

**Background:** Real-world evidence shows alarmingly suboptimal utilization of GDMT in HFrEF. One of the barriers of GDMT implementation appears to be concerns about the potential development of drug-related adverse events (AEs), particularly in high-risk patients. This study aimed to evaluate whether advanced HFrEF patients can be up-titrated safely and whether advanced HFrEF predisposes to the occurrence of putatively drug-related AEs.

**Methods:** A total of 373 HFrEF patients from our prospective HF registry were analyzed for HF drug utilization and target dosages (TDs) at baseline, 2 months, and 12 months. Successful up-titration and AEs were evaluated for different stages of HF reflected by NT-proBNP (<1000pg/ml, 1000-2000pg/ml, >2000pg/ml).

**Results:** A stepwise increase in HF medications could be observed for all drug classes during follow-up. At 12 months 73%, 75%, 62%, 86% and 45% of patients received ≥90% of TDs of beta-blockers (BB), renin-angiotensin system inhibitors (RASi), mineralocorticoid receptor antagonists (MRA), sodium-glucose cotransporter-2 inhibitors (SGLT2i), and triple-therapy, respectively. Predictors of successful up-titration in logistic regression were baseline HF-drug TDs, eGFR, and potassium, but not NT-proBNP or age. The development of AEs was rare, with hyperkalemia as the most common event (34% at 12 months). AEs were comparable in all stages of HF. However, development of hyperkalemia was more frequent in patients with higher NT-proBNP and also accounted for most cases for incomplete up-titration.

**Conclusions:** This study suggests that with dedicated protocols and frequent visits GDMT can be successfully implemented across all stages of HFrEF, including patients with highest NT-proBNP levels who probably profit the most.

**CLINICAL PERSPECTIVE:** *What is new?:*

- GDMT up-titration is feasible even in advanced HFrEF patients, with this study demonstrating that ≥50% target doses for all HF medications can be achieved in about 90% of patients within 12 months through high-intensity care and frequent follow-up visits.
- Adverse events during up-titration are rare, even in high-risk patients, and the most notable AE, hyperkalemia, can be effectively managed in most cases without discontinuing therapy.

*What are the clinical implications?:*

- This study demonstrates that GDMT up-titration is achievable even in advanced HFrEF, challenging the notion that medication intolerance should be part of the definition of advanced HF.
- Practicing physicians should pursue GDMT up-titration for all HFrEF patients, as AEs are manageable, and its potential benefit to stabilize disease progression and improve outcomes far outweigh the risks.
- The lack of consensus on when to halt up-titration or define true intolerance, combined with limited evidence on GDMT efficacy or futility in advanced HF, underscores the need for further research to optimize treatment in this high-risk group.

## INTRODUCTION

Heart failure (HF) remains a leading cause of morbidity and mortality worldwide.^1,2^ Despite transformative advances in HF therapies, their implementation and up-titration in clinical practice lags alarmingly behind. Only 22% of HF patients receive some form of triple therapy, and a mere 1% reach the recommended target doses of all essential medications.^3^

Previously reported barriers of GDMT implementation include therapeutic inertia, cardiology vs non-cardiology care, misperceptions about a “clinically stable” status, potential biases against older, female, or co-morbid patients and concerns over therapy-related adverse events (AEs) as hypotension, impaired renal function or hyperkalemia.^3,4,5,6,7^ Clinicians tend to most frequently underuse GDMT in advanced HF patients, due to a lack of strong evidence in this patient population and concerns about drug-related AEs, while these patients potentially benefit most.^8^ Hypotension, impaired renal function and hyperkalemia are especially common in advanced HFrEF patients, and can be viewed as signs of disease progression as well as drug related AEs while up-titrating HF medications. However, in these patients GDMT might even stabilize the failing heart and kidneys, resulting in more steady blood pressure and renal function. In order not to unnecessarily down-titrate HF drugs and lose out on their benefits, understanding the relationship between disease progression, HF medication use and the development of therapy limiting clinical features would be crucial. It also must be noted, that the cut-offs for defining those HF therapy limiting factors are used inconsistently and are arbitrary by nature.

This study aimed to i) explore whether advanced HFrEF patients can be up-titrated safely with HF medications and ii) assess the development of drug (and disease)-related AEs during medical up-titration in a dedicated HF outpatient unit in a tertiary care center.

## METHODS

### Patient population

Since 2015 a HFrEF registry at the Vienna General Hospital includes systematically chronic HFrEF outpatients defined by a history of documented left ventricular ejection fraction (LVEF) of ≤35% by echocardiography and a history of significantly elevated NT-proBNP levels >1500 pg/ml. The clinic is dedicated to GDMT optimization, device therapy and patient management for advanced HF therapies such as cardiac transplantation or long-term mechanical circulatory support. Clinical data, patient history, cardiac imaging results, medication including dosages as well as laboratory values are routinely documented at first presentation and all subsequent visits. For this study, data of patients between January 2015 and December 2023 were analyzed, who after the baseline visit completed both a short-term (2±1 months, 2M visit) and a long-term follow-up visit (12±6 months, 12M visit). The investigations were conducted in strict adherence to the principles outlined in the Declaration of Helsinki and received institutional ethics committee approval (EK1612/2015). All participants provided written informed consent.

### Assessing GDMT in HF

Precise use and exact dosage of HF medications, i.e. BB, RASi, MRA and SGLT2i were documented at each visit. For comparability, HF medication dosages were expressed as a percentage of the recommended target dosages. Non-receipt of a specific drug was recorded as 0%. Additionally, triple-therapy intensity was assessed by calculating the mean percentage of the TD for BB, RASi, and MRA [*triple* − *therapy TD*% = 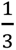 ∗ (BB TD% + RASi TD% + MRA TD%)].^9,10^ SGLT2i data in this paper was only analyzed for patients enrolled after 2021, in alignment with the ESC recommendations for SGLT2i use.

### Definition of clinical factors limiting GDMT up-titration - adverse events

The presence of clinical factors potentially limiting GDMT up-titration / AEs related to HF treatment were assessed at each visit. AEs were defined according to the 2021 ESC HF- guidelines.^9^ The most common clinical AEs were defined as follows: bradycardia, indicated by a resting heart rate (RHR) of <50 beats per minute (bpm); asymptomatic hypotension, identified by an office systolic blood pressure (SBP) of <90 mmHg without symptoms; symptomatic hypotension, identified by an office SBP of <90 mmHg with symptoms; impaired renal function, indicated by an estimated glomerular filtration rate (eGFR) of <30 ml/min/1.73m²; and hyperkalemia (HK), indicated by a serum potassium (K) level of >5.0 mmol/l or >5.5 mmol/l. AEs which were not present as the initial visit but developed at FUP during up-titration were termed as new AEs. In addition, in case that HF drugs were not up-titrated further to TDs beyond 50% medical records were investigated to identify the reason for no up-titration.

### Statistical Analysis

To describe the patient population, baseline characteristics are presented. Continuous data are expressed as median and interquartile range (IQR), categorial data as counts and percentages. To compare the BL characteristics according to up-titrational success, counts were analyzed by the 2-sided Fisher’s exact test, and continuous variables by the Kruskal-Wallis test.

To assess the success of HF drug up-titration during the outpatient visits, target dosages for each drug class as well as triple therapy were compared by the Friedman test across the timepoints (paired, non-parametric test). To identify specific differences between time points, post hoc analysis was performed using pairwise Wilcoxon Signed-Rank Tests (paired, non-parametric test). To correct for multiple comparisons the Bonferroni correction was applied.

For visualization HF drug dosages were summarized in four clinically relevant groups, i.e zero (0%), low (>0%–<50%), medium (≥50%–<90%), and high (≥90%) dosages of TD%.^10^ Changes in HF drug dosage groups were visualized by histograms.

To identify factors associated with up-titrational success i.e. TDs of ≥ 90% at 12-month follow-up, a logistic bootstrap regression model with stepwise forward selection (p ≤ 0.05 for inclusion) as an exploratory method was used. Potential influencing factors considered included: age, sex, BMI, NYHA class, systolic blood pressure (SBP), heart rate (HR), potassium (K), estimated glomerular filtration rate (eGFR), blood urea nitrogen (BUN), sodium, butyrylcholinesterase (BChE), GOT, GPT, GGT, bilirubin, transferrin saturation (TSAT), hemoglobin, triglycerides, C-reactive protein (CRP), NT-proBNP, number of comorbidities, and baseline HF medication dosages. The selection process followed the method described by Harrell.^11^ It accounts for potential non-linear associations and initially excludes variables with weak bivariate correlations (p > 0.2). Variables selected in fewer than 40% of 500 bootstrap samples using forward selection were then excluded step by step. A final logistic regression with forward selection was performed on the remaining predictors. Results are reported in units of standard deviation. Alternative selection methods were tested by using continuous TD values, case-by-case exclusion of missing data or imputation by the mean, and consideration of first-order intercorrelations. These alternatives produced qualitatively consistent results for the key variables (p ≤ 0.01).

To visualize the main results of the logistic regression HF drug dosages in TD% groups were displayed for groups of clinically relevant variables, i.e. NT-proBNP, eGFR, age, sex, BMI and comorbidity burden. In this analysis trends for GDMT up-titration were analyzed between the strata by the Jonckheere-Terpstra test (unpaired, non-parametric test, which calculates p for trend). Sex differences were compared via the Kruskal-Wallis H test (unpaired, non-parametric test).

The prevalence of AEs was displayed as percentages at different timepoints. AEs were compared by the McNemar-Bowker test (paired, categorical test) between timepoints. For an explorative analysis, the distribution of new AEs after 12 months according to subgroups (NT-proBNP, eGFR, age, BMI, sex and number of comorbidities) was compared by the 2- sided Fishers’s exact test (unpaired, categorial test).

IBM SPSS 25.0 and GraphPad Prism 9 were used to perform statistical analyses and create the figures. GChaos 31.3 statistical software written in C++ by one of the authors (GS) was used for the bootstrap selection process. A two-tailed p-value <0.05 was deemed statistically significant. In case of multiple testing, p-values were adjusted by Bonferroni correction.

## RESULTS

### Study population

Baseline characteristics for the total cohort (n=373) are displayed in Table 1. The median age was 62 years (IQR: 50-72), 23.3% of patients were women. 7.8%, 51.6% and 40.6% of patients were NYHA class I, II and III/IV, median NT-proBNP levels were 2363 pg/ml (IQR: 1014-5009). At baseline 93%, 88.7%, 72.7% and 52.8% of patients received BB, RASi, MRA and SGLT2i therapy. 65.1%, 54.2%, and 70.8% of patients were receiving ≥50% of the target dosages, for BB, RASi and MRA, respectively. More severe disease was associated with lower baseline eGFR and hemoglobin levels, higher CRP levels, increased heart rate, higher age, and a greater prevalence of atrial fibrillation. Notably, the use and TDs of BB and SGLT2i were consistent across all NT-proBNP groups. However, patients with NT-proBNP levels >2000 pg/ml had lower usage and TDs of RASi and MRA at baseline.

**Table 1.**
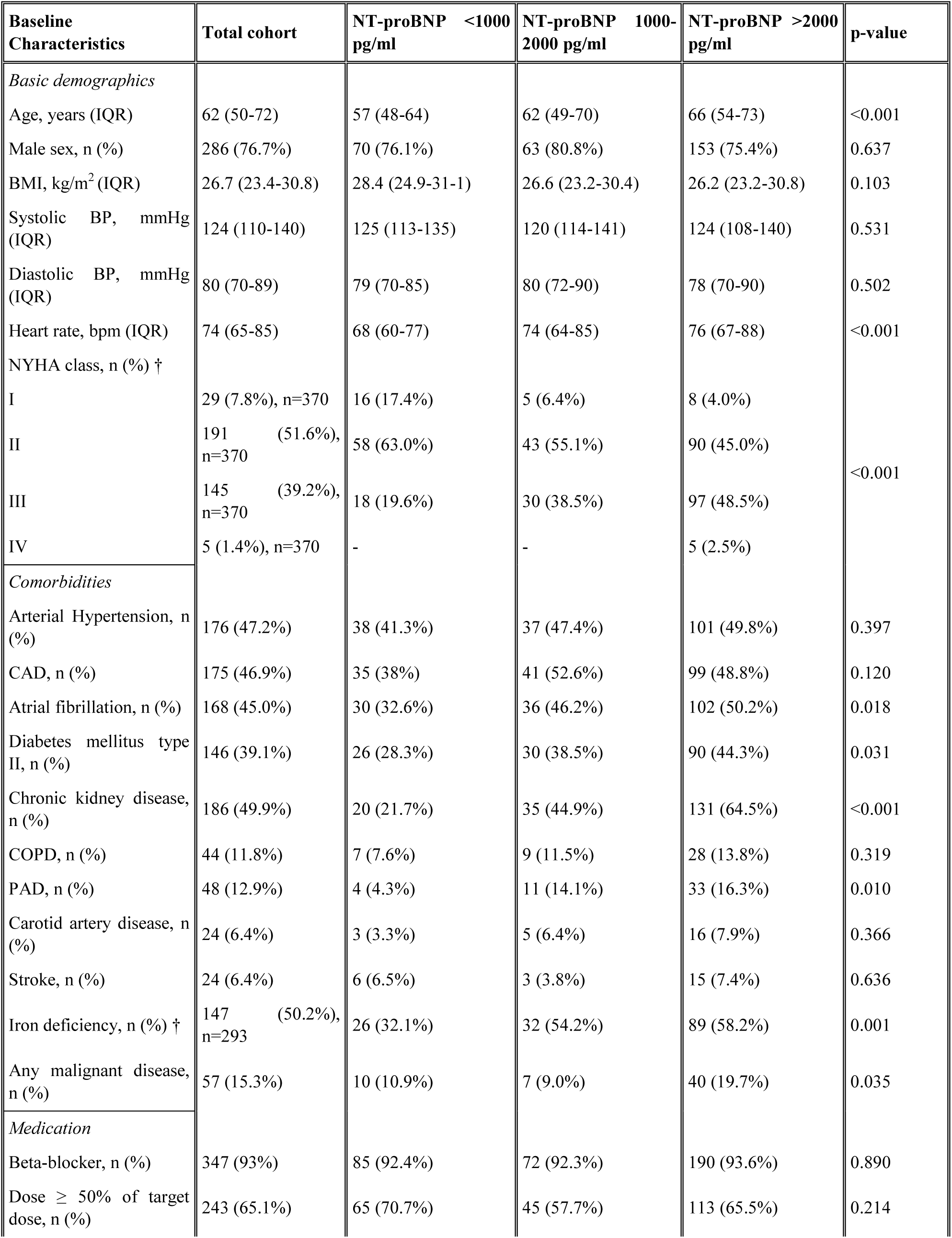
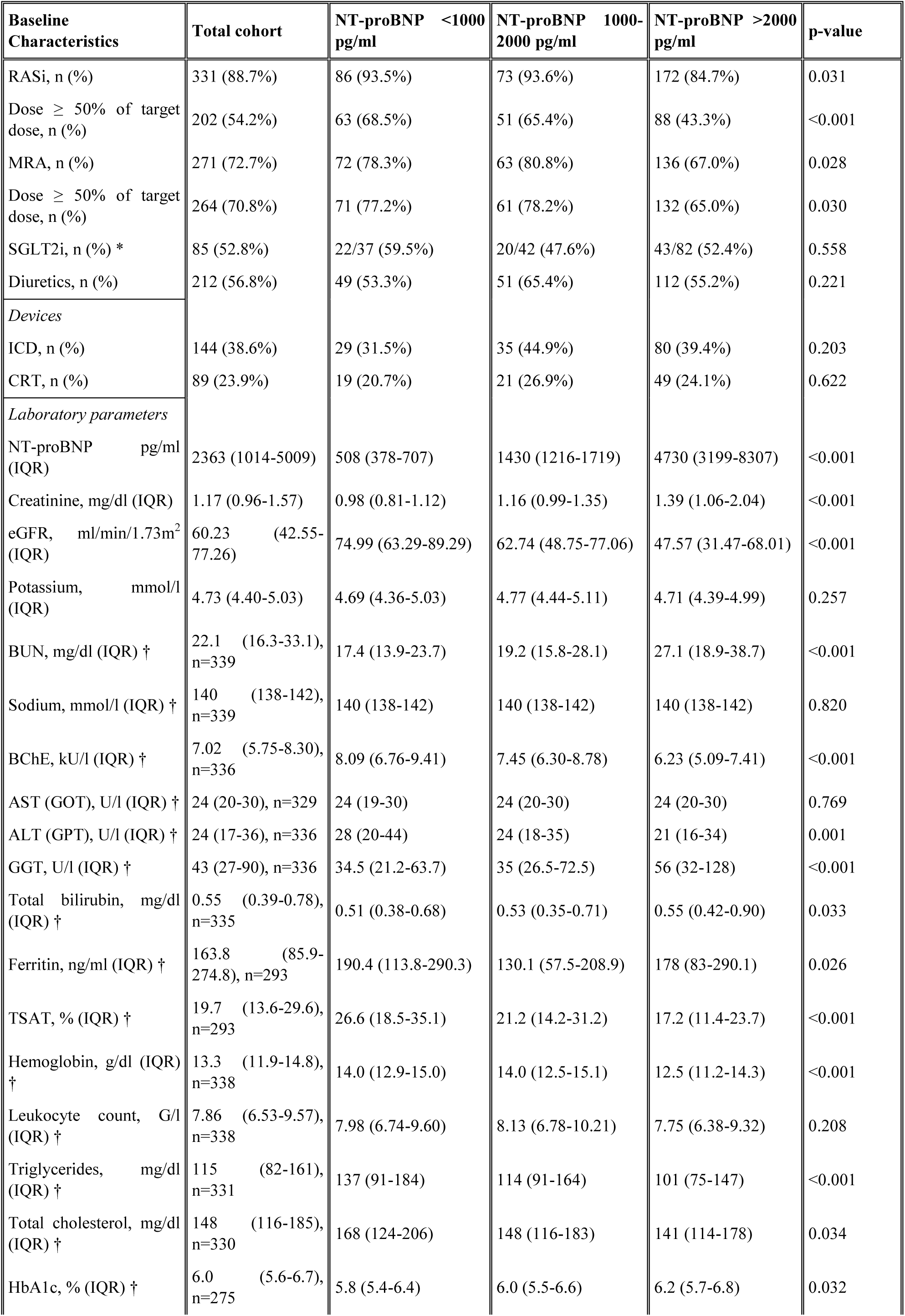
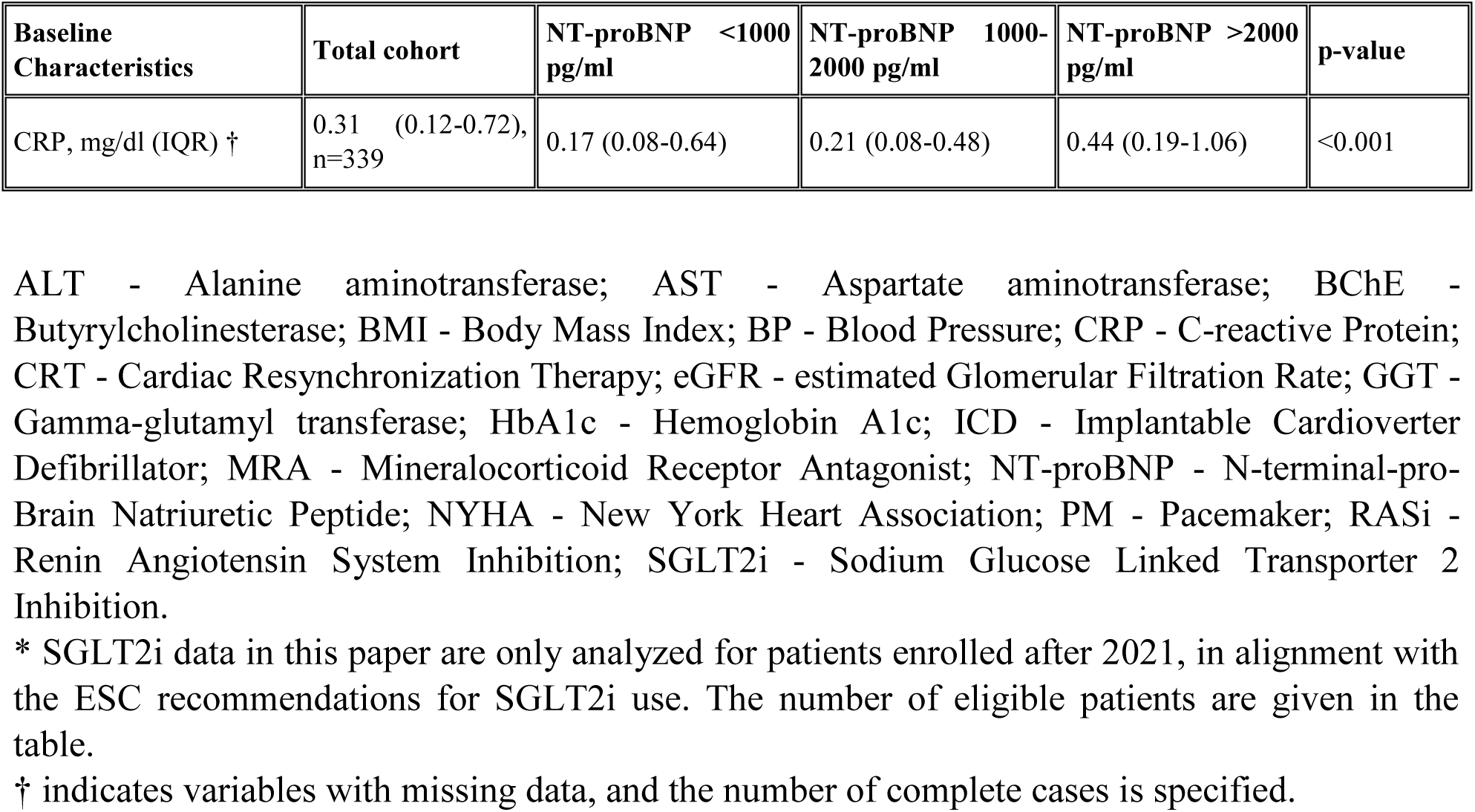
Baseline Characteristics according to NT-proBNP levels.

| Baseline Characteristics | Total cohort | NT-proBNP <1000 pg/ml | NT-proBNP 1000-2000 pg/ml | NT-proBNP >2000 pg/ml | p-value |
| --- | --- | --- | --- | --- | --- |
| Basic demographics |  |  |  |  |  |
| Age, years (IQR) | 62 (50-72) | 57 (48-64) | 62 (49-70) | 66 (54-73) | <0.001 |
| Male sex, n (%) | 286 (76.7%) | 70 (76.1%) | 63 (80.8%) | 153 (75.4%) | 0.637 |
| BMI, kg/m<sup>2</sup> (IQR) | 26.7 (23.4-30.8) | 28.4 (24.9-31.1) | 26.6 (23.2-30.4) | 26.2 (23.2-30.8) | 0.103 |
| Systolic BP, mmHg (IQR) | 124 (110-140) | 125 (113-135) | 120 (114-141) | 124 (108-140) | 0.531 |
| Diastolic BP, mmHg (IQR) | 80 (70-89) | 79 (70-85) | 80 (72-90) | 78 (70-90) | 0.502 |
| Heart rate, bpm (IQR) | 74 (65-85) | 68 (60-77) | 74 (64-85) | 76 (67-88) | <0.001 |
| NYHA class, n (%) † |  |  |  |  |  |
| I | 29 (7.8%), n=370 | 16 (17.4%) | 5 (6.4%) | 8 (4.0%) | <0.001 |
| II | 191 (51.6%), n=370 | 58 (63.0%) | 43 (55.1%) | 90 (45.0%) |  |
| III | 145 (39.2%), n=370 | 18 (19.6%) | 30 (38.5%) | 97 (48.5%) |  |
| IV | 5 (1.4%), n=370 | - | - | 5 (2.5%) |  |
| Comorbidities |  |  |  |  |  |
| Arterial Hypertension, n (%) | 176 (47.2%) | 38 (41.3%) | 37 (47.4%) | 101 (49.8%) | 0.397 |
| CAD, n (%) | 175 (46.9%) | 35 (38%) | 41 (52.6%) | 99 (48.8%) | 0.120 |
| Atrial fibrillation, n (%) | 168 (45.0%) | 30 (32.6%) | 36 (46.2%) | 102 (50.2%) | 0.018 |
| Diabetes mellitus type II, n (%) | 146 (39.1%) | 26 (28.3%) | 30 (38.5%) | 90 (44.3%) | 0.031 |
| Chronic kidney disease, n (%) | 186 (49.9%) | 20 (21.7%) | 35 (44.9%) | 131 (64.5%) | <0.001 |
| COPD, n (%) | 44 (11.8%) | 7 (7.6%) | 9 (11.5%) | 28 (13.8%) | 0.319 |
| PAD, n (%) | 48 (12.9%) | 4 (4.3%) | 11 (14.1%) | 33 (16.3%) | 0.010 |
| Carotid artery disease, n (%) | 24 (6.4%) | 3 (3.3%) | 5 (6.4%) | 16 (7.9%) | 0.366 |
| Stroke, n (%) | 24 (6.4%) | 6 (6.5%) | 3 (3.8%) | 15 (7.4%) | 0.636 |
| Iron deficiency, n (%) † | 147 (50.2%), n=293 | 26 (32.1%) | 32 (54.2%) | 89 (58.2%) | 0.001 |
| Any malignant disease, n (%) | 57 (15.3%) | 10 (10.9%) | 7 (9.0%) | 40 (19.7%) | 0.035 |
| Medication |  |  |  |  |  |
| Beta-blocker, n (%) | 347 (93%) | 85 (92.4%) | 72 (92.3%) | 190 (93.6%) | 0.890 |
| Dose ≥ 50% of target dose, n (%) | 243 (65.1%) | 65 (70.7%) | 45 (57.7%) | 113 (65.5%) | 0.214 |
| RASi, n (%) | 331 (88.7%) | 86 (93.5%) | 73 (93.6%) | 172 (84.7%) | 0.031 |
| Dose ≥ 50% of target dose, n (%) | 202 (54.2%) | 63 (68.5%) | 51 (65.4%) | 88 (43.3%) | <0.001 |
| MRA, n (%) | 271 (72.7%) | 72 (78.3%) | 63 (80.8%) | 136 (67.0%) | 0.028 |
| Dose ≥ 50% of target dose, n (%) | 264 (70.8%) | 71 (77.2%) | 61 (78.2%) | 132 (65.0%) | 0.030 |
| SGLT2i, n (%) * | 85 (52.8%) | 22/37 (59.5%) | 20/42 (47.6%) | 43/82 (52.4%) | 0.558 |
| Diuretics, n (%) | 212 (56.8%) | 49 (53.3%) | 51 (65.4%) | 112 (55.2%) | 0.221 |
| *Devices* |  |  |  |  |  |
| ICD, n (%) | 144 (38.6%) | 29 (31.5%) | 35 (44.9%) | 80 (39.4%) | 0.203 |
| CRT, n (%) | 89 (23.9%) | 19 (20.7%) | 21 (26.9%) | 49 (24.1%) | 0.622 |
| *Laboratory parameters* |  |  |  |  |  |
| NT-proBNP pg/ml (IQR) | 2363 (1014-5009) | 508 (378-707) | 1430 (1216-1719) | 4730 (3199-8307) | <0.001 |
| Creatinine, mg/dl (IQR) | 1.17 (0.96-1.57) | 0.98 (0.81-1.12) | 1.16 (0.99-1.35) | 1.39 (1.06-2.04) | <0.001 |
| eGFR, ml/min/1.73m<sup>2</sup> (IQR) | 60.23 (42.55-77.26) | 74.99 (63.29-89.29) | 62.74 (48.75-77.06) | 47.57 (31.47-68.01) | <0.001 |
| Potassium, mmol/l (IQR) | 4.73 (4.40-5.03) | 4.69 (4.36-5.03) | 4.77 (4.44-5.11) | 4.71 (4.39-4.99) | 0.257 |
| BUN, mg/dl (IQR) † | 22.1 (16.3-33.1), n=339 | 17.4 (13.9-23.7) | 19.2 (15.8-28.1) | 27.1 (18.9-38.7) | <0.001 |
| Sodium, mmol/l (IQR) † | 140 (138-142), n=339 | 140 (138-142) | 140 (138-142) | 140 (138-142) | 0.820 |
| BChE, kU/l (IQR) † | 7.02 (5.75-8.30), n=336 | 8.09 (6.76-9.41) | 7.45 (6.30-8.78) | 6.23 (5.09-7.41) | <0.001 |
| AST (GOT), U/l (IQR) † | 24 (20-30), n=329 | 24 (19-30) | 24 (20-30) | 24 (20-30) | 0.769 |
| ALT (GPT), U/l (IQR) † | 24 (17-36), n=336 | 28 (20-44) | 24 (18-35) | 21 (16-34) | 0.001 |
| GGT, U/l (IQR) † | 43 (27-90), n=336 | 34.5 (21.2-63.7) | 35 (26.5-72.5) | 56 (32-128) | <0.001 |
| Total bilirubin, mg/dl (IQR) † | 0.55 (0.39-0.78), n=335 | 0.51 (0.38-0.68) | 0.53 (0.35-0.71) | 0.55 (0.42-0.90) | 0.033 |
| Ferritin, ng/ml (IQR) † | 163.8 (85.9-274.8), n=293 | 190.4 (113.8-290.3) | 130.1 (57.5-208.9) | 178 (83-290.1) | 0.026 |
| TSAT, % (IQR) † | 19.7 (13.6-29.6), n=293 | 26.6 (18.5-35.1) | 21.2 (14.2-31.2) | 17.2 (11.4-23.7) | <0.001 |
| Hemoglobin, g/dl (IQR) † | 13.3 (11.9-14.8), n=338 | 14.0 (12.9-15.0) | 14.0 (12.5-15.1) | 12.5 (11.2-14.3) | <0.001 |
| Leukocyte count, G/l (IQR) † | 7.86 (6.53-9.57), n=338 | 7.98 (6.74-9.60) | 8.13 (6.78-10.21) | 7.75 (6.38-9.32) | 0.208 |
| Triglycerides, mg/dl (IQR) † | 115 (82-161), n=331 | 137 (91-184) | 114 (91-164) | 101 (75-147) | <0.001 |
| Total cholesterol, mg/dl (IQR) † | 148 (116-185), n=330 | 168 (124-206) | 148 (116-183) | 141 (114-178) | 0.034 |
| HbA1c, % (IQR) † | 6.0 (5.6-6.7), n=275 | 5.8 (5.4-6.4) | 6.0 (5.5-6.6) | 6.2 (5.7-6.8) | 0.032 |
| CRP, mg/dl (IQR) † | 0.31 (0.12-0.72), n=339 | 0.17 (0.08-0.64) | 0.21 (0.08-0.48) | 0.44 (0.19-1.06) | <0.001 |

ALT - Alanine aminotransferase; AST - Aspartate aminotransferase; BChE - Butyrylcholinesterase; BMI - Body Mass Index; BP - Blood Pressure; CRP - C-reactive Protein; CRT - Cardiac Resynchronization Therapy; eGFR - estimated Glomerular Filtration Rate; GGT - Gamma-glutamyl transferase; HbA1c - Hemoglobin A1c; ICD - Implantable Cardioverter Defibrillator; MRA - Mineralocorticoid Receptor Antagonist; NT-proBNP - N-terminal-pro-Brain Natriuretic Peptide; NYHA - New York Heart Association; PM - Pacemaker; RASi - Renin Angiotensin System Inhibition; SGLT2i - Sodium Glucose Linked Transporter 2 Inhibition.
\* SGLT2i data in this paper are only analyzed for patients enrolled after 2021, in alignment with the ESC recommendations for SGLT2i use. The number of eligible patients are given in the table.
† indicates variables with missing data, and the number of complete cases is specified.

### Up-titration of HF medication

The achieved TDs for all HF drug classes at BL, 2M and 12M are shown in Figure 1. A significant increase in TDs could be observed for all drug classes during FUP (p<0.001 for comparisons between BL vs 2M and BL vs 12M for the continuous variable of TDs). The majority of patients received ≥50% and ≥90% of the recommended TDs at 12M (for ≥50%: 90%, 90%, 84%, 86% and 91%; and for ≥90%: 73%, 75%, 62%, 86% and 45% for BB, RASi, MRA, SGLT2i and triple-therapy, respectively).

**Figure 1.**
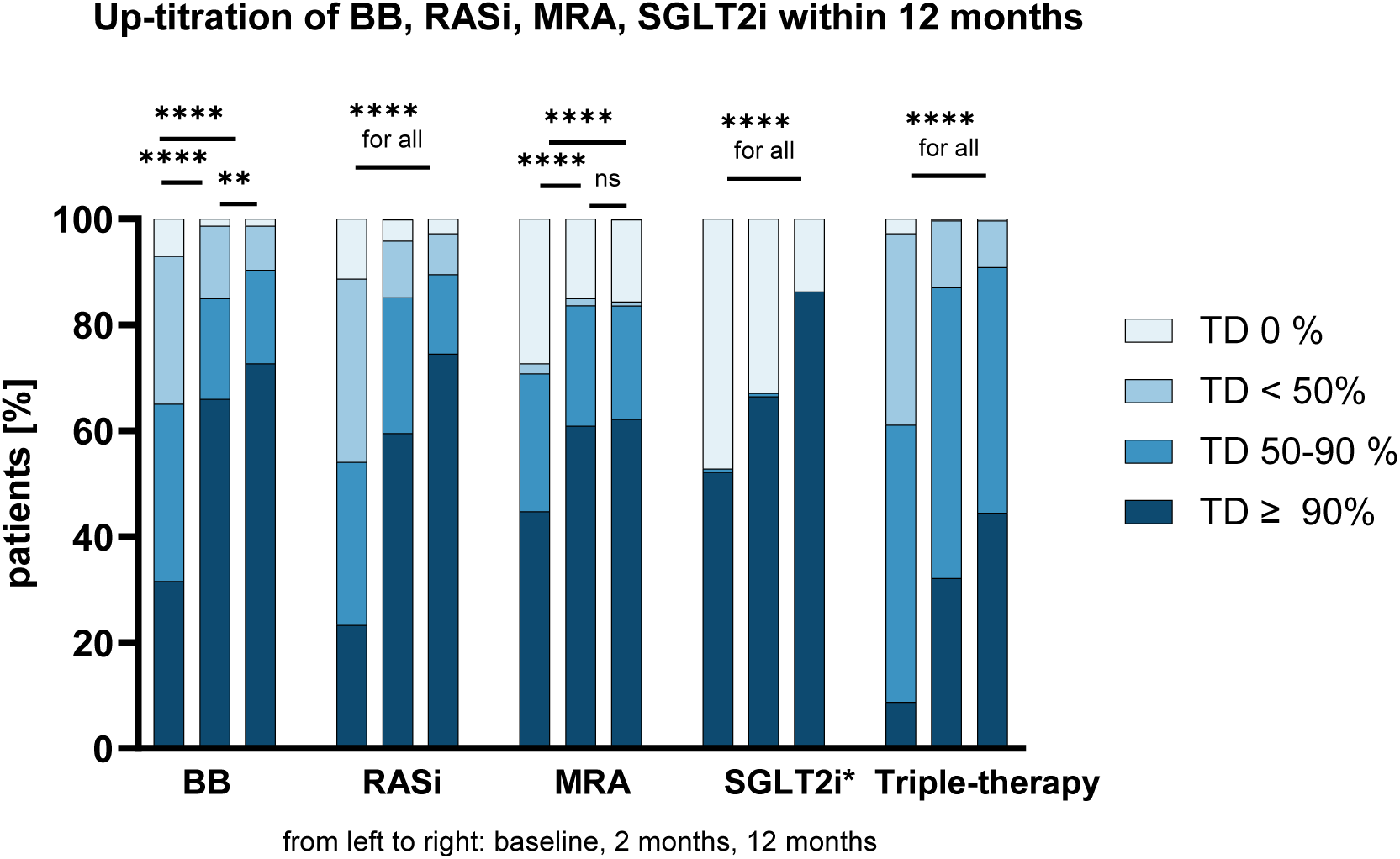
Up-titration of HF medication in a HFrEF cohort at a tertiary care center within 12 months. The percentage of patients with the achieved target dose (TD) of betablockers (BB), renin-angiotensin-system inhibitors (RASi), mineralocorticoid receptor antagonists (MRA), sodium–glucose co-transporter 2 inhibitors (SGLT2i*, since 2021) and triple-therapy are shown for the total cohort at baseline, 2 months and 12 months. Data was compared by the Friedman test and post hoc analysis using Bonferroni correction. ns=not significant; * p<0.05; ** p<0.01; *** p<0.001; **** p<0.0001

### Predictors for successful up-titration– impact of disease severity and patient characteristics

A multiple linear regression was conducted to determine the predictors of achieved TD of triple-therapy at 12 months. The overall model was statistically significant (R^2^=0.150, Cox & Snell). Significant predictors at baseline were eGFR (ß=0.679, p<0.001), triple-therapy (ß=0.523, p<0.001) and serum potassium (ß=-0.240, p=0.048). Up-titrational success was especially independent from heart failure severity reflected by NT-proBNP levels or NYHA class, but also age, comorbidity burden, sex and BMI. Figure 2 displays the main findings graphically. There was no consistent difference in successful up-titration for NT-proBNP, age, sex or BMI. However, a consistent trend for lower GDMT was observable with increasingly impaired renal function (BB: p<0.001, RASi: p=0.001, MRA: p<0.001, SGLT2i: p=ns, triple-therapy: p<0.001).

**Figure 2.**
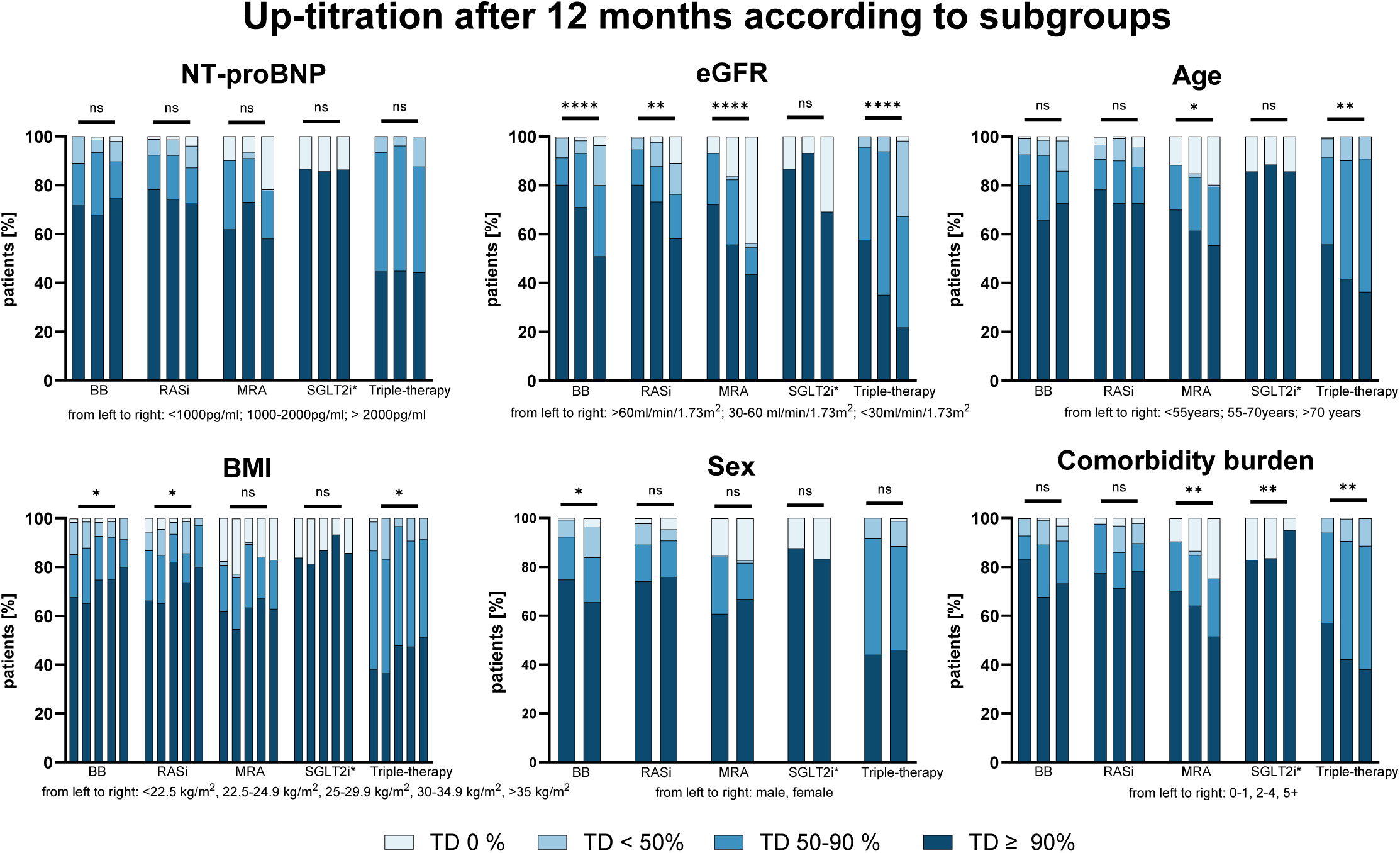
Up-titration of HF medication according to subgroups after 12 months. The proportion of patients receiving the achieved % of the recommended TDs of betablockers (BB), renin-angiotensin-system inhibitors (RASi), mineralocorticoid receptor antagonists (MRA), sodium– glucose co-transporter 2 inhibitors (SGLT2i*, since 2021) and triple-therapy are shown after 12M according to NT-proBNP, eGFR, age, BMI, sex and comorbidity burden. Data trends were analyzed by the Jonckheere-Terpstra test, sex was evaluated by the Kruskal-Wallis H test. ns=not significant; * p<0.05; ** p<0.01; *** p<0.001; **** p<0.0001

### Frequency of adverse events – impact of disease severity and patient characteristics

Figure 3 shows the prevalence and development of AEs at BL, 2M and 12M. At baseline 3% of patients exhibited hypotension, 2.4% bradycardia, 14.7% impaired renal function and 25.5% mild (K >5.0 mmol/l) and 7.8% significant (K >5.5 mmol/l) hyperkalemia. The development of new AEs was generally infrequent, with less than 5% of cases experiencing hypotension, bradycardia, and renal impairment at 2M and 12M. The prevalence of hypotension, bradycardia and impaired renal function remained similar during medical up-titration. New cases of hyperkalemia were the most frequent new AE and developed in 22.9% and 22.8% (K >5.0 mmol/l) and 8.9% and 6.7% (K >5.5 mmol/l) of cases after 2 and 12 months. Figure 4 shows an explorative analysis for the development of HF drug related AEs at 12M for clinically relevant patient subgroups. New AEs as hypotension, bradycardia or impaired renal function were equally distributed across NT-proBNP strata, sex or BMI (p=ns for all). Only the development of hyperkalemia (K >5.0 mmol/l at 12M) increased with higher NT-proBNP (15.2% vs 19.2% vs 27.6%, p=0.045). Generally, the development of hyperkalemia (K >5.0 mmol/l at 12M) and impaired renal function (drop of eGFR below <30ml/min/1.73m^2^) was associated with higher age and increased comorbidity burden. Both higher age and increased comorbidity burden predispose for worse baseline renal function.

**Figure 3.**
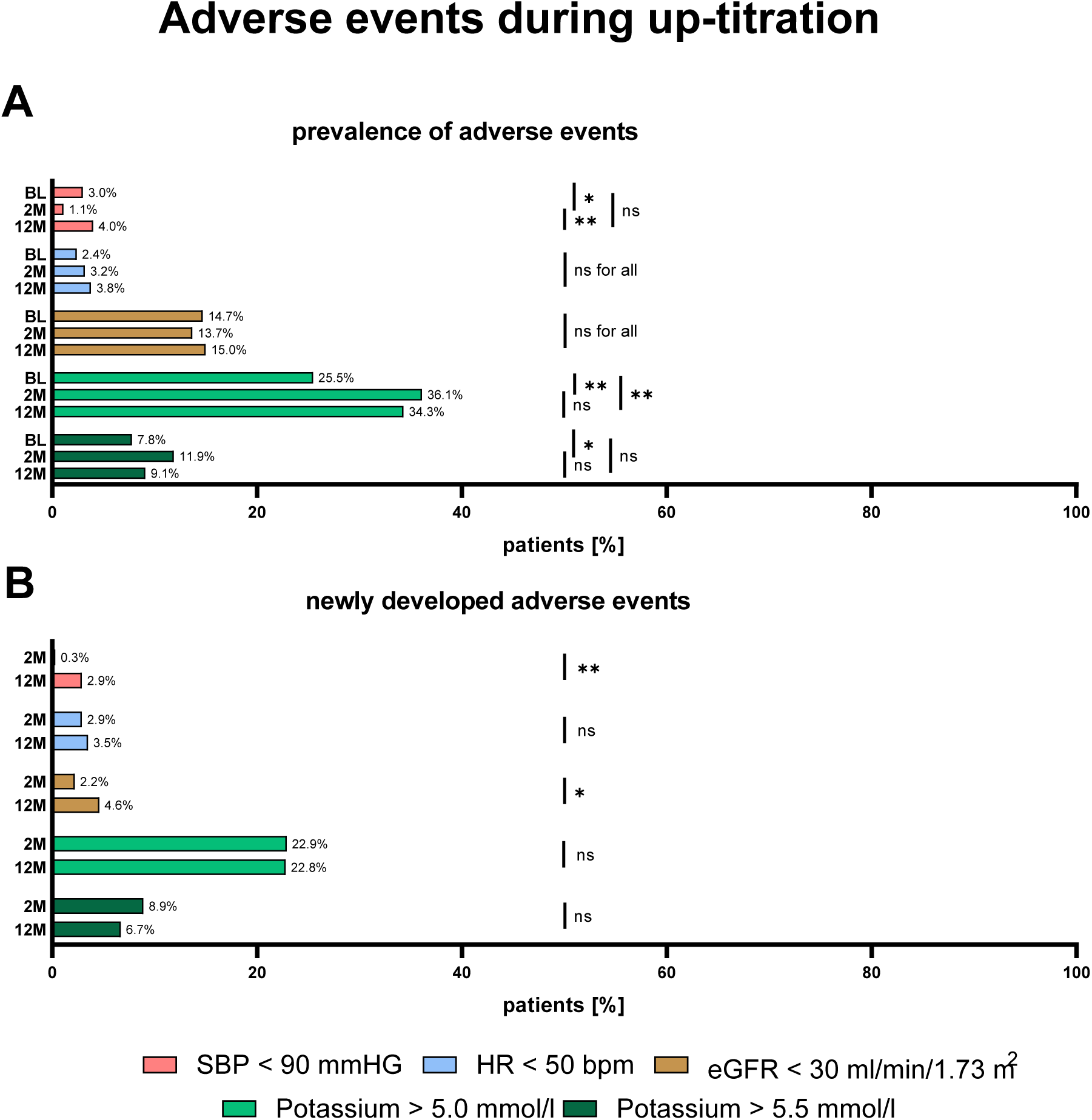
Prevalence of adverse events (AEs) and newly development AEs. The percentage of patients who developed AEs at baseline (BL), 2 months (2M) or 12 months (12M) (A) and percentage of patients with newly developed AEs (B) at 2M or 12M are shown. Depicted is the percentage of patients experiencing systolic blood pressure (SBP) <90 mmHG, heart rate (HR) <50 beats per minute (bpm), estimated glomerular filtration rate (eGFR) <30 ml/min/1.73 m^2^, potassium (K) >5.0 mmol/l and K >5.5 mmol/l. Data was compared by the McNemar-Bowker test. ns=not significant; * p<0.05; ** p<0.01; *** p<0.001; **** p<0.0001

**Figure 4.**
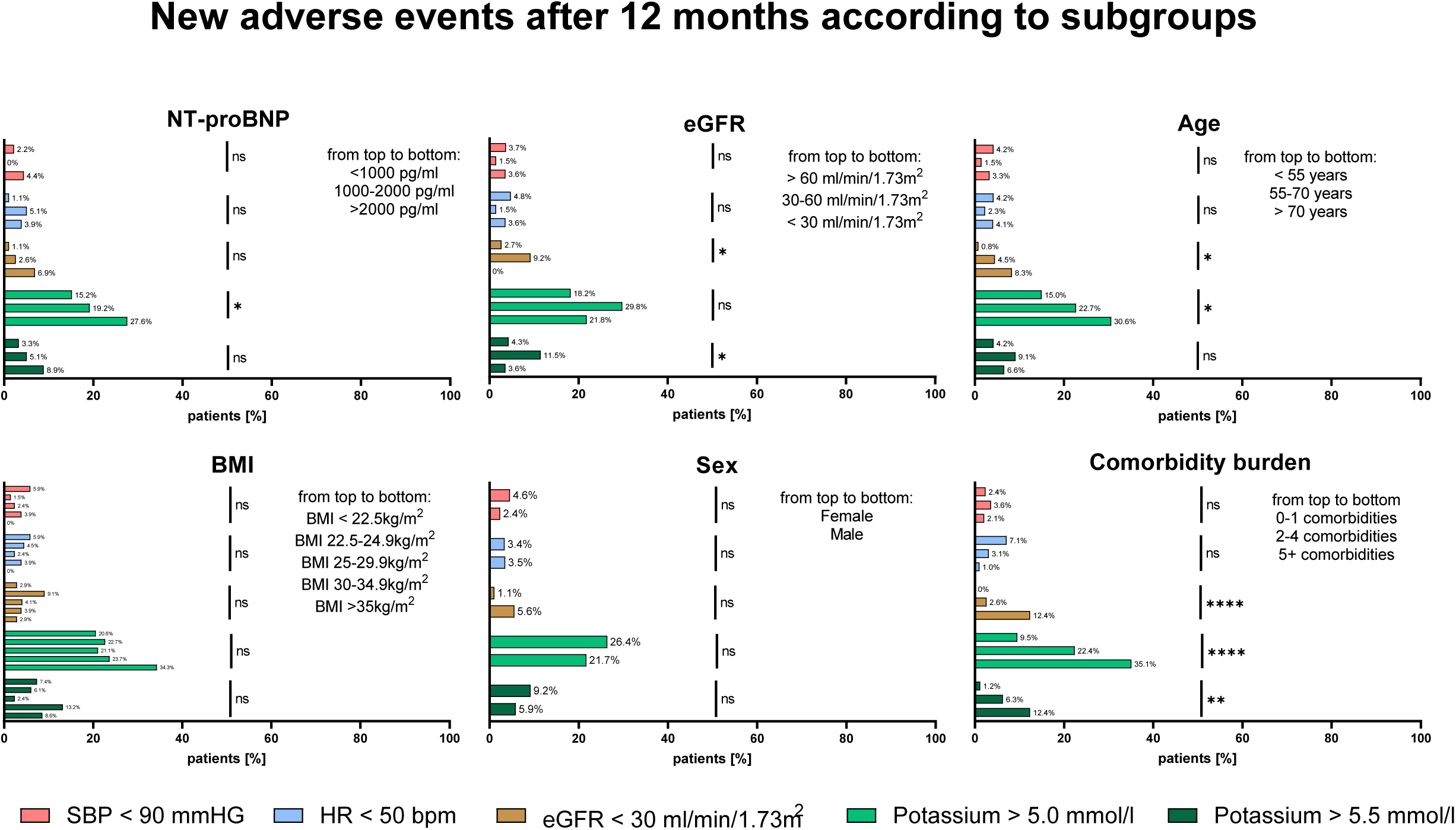
Distribution of newly developed adverse events (AEs) after 12 months according to subgroups. The percentage of patients who developed new AEs at 12 M according to NT-proBNP, eGFR, age, BMI, sex and comorbidity burden are shown. Depicted is the percentage of patients experiencing systolic blood pressure (SBP) <90 mmHG, heart rate (HR) <50 beats per minute (bpm), estimated glomerular filtration rate (eGFR) <30 ml/min/1.73 m2, potassium (K) >5.0 mmol/l and K >5.5mmol/l. Data was compared by the Fishers’s exact test. ns=not significant; * p<0.05; ** p<0.01; *** p<0.001; **** p<0.0001

### Reasons for no up-titration (TD ≤ 50%) at 12M

Table 2 shows the reason for maintaining suboptimal TDs. After 12 months, 23%, 22%, and 38% of patients could not be up-titrated beyond 50% of the recommended target dose. Likely causes could be identified for 92%, 91% and 75% of BB, RASi, and MRA cases. The most common causes were bradycardia and hypotension for BBs (in total accounting for 66%), hypotension and hyperkalemia for RASi (in total accounting for 61%), and hyperkalemia with or without impaired renal function for MRAs (in total accounting for 58%). Among all HFrEF medications, MRAs were most likely to remain at suboptimal dosages and showed the highest number of cases where no reason could be identified. Hyperkalemia accounted for a total of 31% and 43% of cases for suboptimal dosages of RASi and MRA.

**Table 2.** Logistic regression model for up-titrational success at 12 months.

| Variable | β-Coefficient | Standard Error | T-value | p-value | Odds Ratio | 95% CI | Cox & Snell Pseudo $R^2$ |
| --- | --- | --- | --- | --- | --- | --- | --- |
| Intercept | -0.298 |  |  |  |  |  | 0.150 |
| eGFR (baseline)* | 0.679 | 0.143 | 4.733 | <0.001 | 1.972 | 1.488-2.612 |  |
| Triple-therapy TD (baseline)* | 0.523 | 0.118 | 4.432 | <0.001 | 1.686 | 1.338-2.125 |  |
| Serum potassium (baseline)* | -0.240 | 0.121 | 1.975 | 0.048 | 0.787 | 0.620-0.998 |  |

\* Variables are z-transformed, increase as per standard unit

**Table 3.** Reasons for suboptimal GDMT (TD ≤ 50%) at 12 months.

| TD GDMT ≤ 50% | BB TD ≤ 50%<br>(n=86/373) | RASi TD ≤ 50%<br>(n=84/373) | MRA TD ≤ 50%<br>(n=141/373) |
| --- | --- | --- | --- |
| *Reasons* |  |  |  |
| asymptomatic hypotension | 16 | 17 | - |
| bradycardia | 32 | - | - |
| hyperkalemia | - | 26 | 61 |
| Impaired renal function | - | 8 | 12 |
| hyperkalemia and Impaired renal function | - | 6 | 21 |
| dizziness | 3 | 1 | - |
| symptomatic hypotension | 9 | 8 | - |
| symptomatic bradycardia | 3 | - | - |
| syncope/presyncope | 2 | - | - |
| intolerance/patient request/allergy | 0/6/0 | 0/3/0 | 0/6/0 |
| medication discontinued elsewhere | 8 | 7 | 5 |
| Total number of reasons | 79/86 (91.9%) | 76/84 (90.5%) | 105/141 (74.5%) |
| no reason | 7/86 (8.1%) | 8 (9.5%) | 36 (25.5%) |

## DISCUSSION

This is the first study to investigate differences in GDMT implementation according to the severity of heart failure and to deliver data on the development of AEs during up-titration of GDMT in advanced HFrEF in a tertiary care outpatient HF center. GDMT therapy implementation was good, whereby 91% of patients achieved ≥50% of the TDs in all recommended drug classes at 1-year. GDMT up-titration could be successfully achieved regardless of NT-proBNP strata or sex. Despite successful up-titration, new AEs remained rare, with only 4%, 4%, 5% and 7% of patients developing bradycardia, hypotension, severe renal impairment (eGFR <30ml/min/1.73m^2^) and severe hyperkalemia (K >5.5 mmol/L). A trend for increasing number of AEs and lower average dosages in GDMT was observed in patients with older age, worse baseline renal function and higher comorbidity burden.

### GDMT evidence in advanced HF

In a recent Universal Definition of heart failure from multiple HF societies advanced HF failure is defined by several key characteristics, such as severe symptoms or symptoms at rest, recurrent HFH despite GDMT, and requirement of advanced therapies such as mechanical circulatory support.^12^ Most importantly, also intolerance to up-titration of GDMT is part of this definition.

Other definitions as the updated HFA-ESC definition, refer to symptoms, NT-proBNP, cardiac dysfunction, but do not mention the inability of up-titration.^13^

There is no established consensus on when to halt GDMT up-titration or how to accurately define true medication intolerance. Moreover, evidence on the efficacy or futility of HF medications in patients with advanced HF is scarce, leaving clinicians with limited guidance for optimizing treatment in this high-risk population.

Trials in high-risk, advanced HF patients, such as CONSENSUS, COPERNICUS, and RALES, demonstrated the efficacy of ACEi, BBs, and MRAs in this population, with significant reductions in mortality and the combined risk of death or HF hospitalization.^14,15,16^ Similarly, in PARADIGM-HF, ARNI showed consistent efficacy regardless of NT-proBNP levels, including those above and below the median of 1631 pg/ml, although patients in general were lower risk.^17^

### GDMT implementation and clinical patient profiles in advanced HFrEF

Global registries reveal suboptimal use of GDMT in HFrEF. In CHAMP-HF, only 28%, 17%, 14%, and 77% of patients achieved TDs for BB, ACEi/ARB, ARNI, and MRA, with just 1.1% on triple therapy at TD.^3^ European data from CHECK-HF show similar results.^18^ While studies focusing on up-titration in stable HFrEF such as GUIDE-IT and BIOSTAT-CHF confirm poor up-titration even after follow-up, the STRONG-HF study demonstrates better results after acute heart failure.^10,19,20,21^

The HFrEF population in this study corresponds to advanced HF and with a median NT-proBNP of 2363 pg/ml [1014-5009] comparable to the population of CHAMP-HF or GUIDE-IT, but also includes patients with lower or intermediate risk.

Up-titration of all four pillars of GDMT was successfully achieved across all heart failure risk groups. TDs were consistent in the low-risk group with NT-proBNP levels <1000 pg/ml (median 508 pg/ml [378-707]), the intermediate-risk group with NT-proBNP levels between 1000-2000 pg/ml (median 1430 pg/ml [1216-1719]), and the high-risk group with NT-proBNP levels >2000 pg/ml (median 4740 pg/ml [3199-8397]), representing the advanced heart failure cohort. Although there is no exact cut-point of NT-proBNP to define advanced heart failure it is noteworthy that patients with advanced HF treated with a ventricular assist device typically have a median NT-proBNP level of about 3500 pg/ml.^22^

### Patient characteristics predisposing for suboptimal GDMT

Patients with more severe heart failure, greater symptoms, or comorbidities often receive lower doses of GDMT. Studies consistently show that factors like older age, higher NYHA-class, greater NT-proBNP levels, low blood pressure, female sex, poor kidney function, and the presence of other comorbidities hold the physicians off up-titration of GDMT.^3,5,18,19,23^ In the CHAMP-HF study, even after 12 months follow-up, dose increases occurred in only 10%, 9%, and 6% of patients for these medications, while 7%, 7%, and 4% had dose reductions.^24^

In the present dataset no bias regarding GDMT implementation based on sex could be observed. Most importantly, GDMT implementation was comparably successful between NT-proBNP strata, indicating that up-titration is feasible and worth pursuing in most severe disease.

In line with previous reports beside eGFR, older age, lower BMI and high comorbidity burden were indeed associated with somewhat worse implementation of GDMT, in univariate analysis. In multivariate analysis kidney function and potassium remained significant, indicating that the influence of age, BMI and comorbidities is largely mediated through eGFR.

Notably, in a logistic regression model NT-proBNP did not predict up-titration at follow-up.

### Development of adverse events during up-titration

The presumably largest limiting factor for GDMT implementation is the expectation of development of HF drug-related AEs. However, data on AEs during up-titration in advanced HF remains scarce, and factors limiting GDMT are often underreported or poorly documented in studies and registries. Notably, the achievement of high TDs in registries is far less common than observed in this study. In BIOSTAT-CHF drug intolerance was cited as a reason for not achieving target doses in 22% of BB patients and 26% of ACEi/ARB, though in most cases, the reasons were unknown or unrecorded.^19^ A secondary analysis of GUIDE-IT found that common reasons for avoiding up-titration included the perception of “clinically stable” status or being “already at maximally tolerated therapy”.^5^ GUIDE-IT reported an overall low rate of symptomatic hypotension (2%), symptomatic bradycardia (0%), hyperkalemia (2.5%), and worsening renal function (3.6%), but did not differentiate between disease severity.^20^

A recent meta-analysis highlighted the misperception of the development of AEs related to HF therapy. The study included landmark cardiovascular outcome trials with forced up-titration in HFrEF and investigated the occurrence of AEs between different HF drugs and placebo.^25^ Almost all clinically relevant AEs were rare and occurred at similar rates in both treatment and control groups. This indicates that events such as drops in blood pressure, worsening renal function, or hyperkalemia are not primarily attributable to specific therapies but probably rather reflect the underlying risks associated with HF itself. The overall frequency of drug-related AEs in these trials was low.^25^ It can be presumed that these underlying risks are a function of disease stages.

This study reinforces these findings showing that the development of HF drug-attributable AEs during medical up-titration is low and that up-titration can be achieved in a dedicated setting. Moreover, it contributes new insights into the tolerability of GDMT and the occurrence of AEs across different risk groups, especially advanced HF and other HF subpopulations. Our analysis identified two key factors as particularly relevant for GDMT up-titration: hyperkalemia and impaired kidney function, both of which were more common in high-risk patients. Notably, higher baseline eGFR and lower potassium levels emerged as strong predictors of successful up-titration. Importantly, modern evidence suggests that neither impaired kidney function nor HK should be considered insurmountable barriers to GDMT optimization, as both conditions can often be effectively managed.

HK is a common issue in heart failure, linked to use of RASi and MRA, as well as severe HF, renal impairment, diabetes, and older age.^26^ However, HK is now manageable with potassium-binding agents, enabling higher doses of RASi and MRAs.^27,28^ Poor kidney function should not preclude GDMT, as studies like STOP-ACEi and the Swedish HF Registry show continuing RASi and MRAs is safe, even in severe renal dysfunction.^29,30^ Moreover, therapies like SGLT2i and ARNI stabilize renal function long-term.^31,32^ Additionally, a recent study demonstrated benefits of ARNI treatment even in patients with end stage renal disease requiring dialysis.^33^ These findings highlight the importance of maintaining GDMT despite HK or renal concerns to optimize outcomes in high-risk HF patients.

Our findings suggest that up-titration of HF medications is feasible and safe across a wide range of NT-proBNP levels. Although GDMT implementation was somewhat lower in patients with older age, higher comorbidity burden and more impaired kidney function, the majority of patients received ≥50% and ≥90% of the recommended TDs at 12M. This underscores that proactive, intensive management should be prioritized for high-risk patients to potentially improve their prognosis.

## Conclusions

Further research is needed to establish stronger evidence on the safety, tolerability, and benefits of GDMT up-titration in advanced HF patients. This study demonstrates that, with dedicated protocols and close follow-up through frequent visits, GDMT can be successfully implemented at much higher rates than reported in previous registries, especially among high-risk, advanced HF patients. Importantly, AEs are generally infrequent and often not directly attributable to HF medications. These findings emphasize that AEs, though more common in high-risk patients, should not deter GDMT up-titration, as its potential to stabilize disease progression and improve outcomes remains significant.

## Limitations

This study has several limitations. As our prospective HFrEF registry focuses on advanced HF patients, the findings may not be generalizable to low-risk populations, though low-risk patients with baseline NT-proBNP levels <1000 pg/ml were included. The specialized HF outpatient setting may not reflect non-specialized care, limiting applicability to primary care or community hospitals. Potential confounders, such as patient adherence, patient requests, and multidisciplinary management, could influence treatment consistency and up-titration decisions. While the study emphasized GDMT up-titration and AEs, other critical factors like quality of life, functional improvement, and risk reduction warrant consideration. Additionally, evolving therapies, such as SGLT2i, Vericiguat, and potassium-binders, were gradually implemented in our patient cohort, which may have impacted GDMT implementation and related factors during the study period (2015–2023).

## Data Availability

Data will not be shared or will be available upon request.

## Abbreviations

ACEi: Angiotensin-Converting Enzyme Inhibitor
ARB: Angiotensin Receptor Blocker
ARNI: Angiotensin Receptor-Neprilysin Inhibitor
BB: Beta Blocker
eGFR: Estimated Glomerular Filtration Rate
GDMT: Guideline Directed Medical Therapy
HF: Heart Failure
HFA: Heart Failure Association
HFrEF: Heart Failure with reduced Ejection Fraction
HK: Hyperkalemia
IQR: Interquartile Range
LVEF: Left Ventricular Ejection Fraction
MRA: Mineralocorticoid Receptor Antagonist
NYHA: New York Heart Association
RASi: Renin Angiotensin System Inhibitor
RHR: Resting Heart Rate
SGLT2i: Sodium-Glucose Transporter 2 Inhibitor

## Acknowledgements

I acknowledge the use of ChatGPT (https://chatgpt.com/) exclusively for text refinement purposes. In certain instances, text passages written by the authors were adapted or condensed with the assistance of ChatGPT. All generated text was thoroughly reviewed and revised by the authors to ensure accuracy and alignment with the manuscript’s intent.

## Sources of Funding

None.

## Disclosures

The authors have nothing to declare.

## Notes

### Competing Interest Statement

The authors have declared no competing interest.

### Funding Statement

No external funding was received

### Author Declarations

All investigations were conducted in strict adherence to the principles outlined in the Declaration of Helsinki and received institutional ethics committee approval (Ethikkommission der Medizinischen Universität Wien, EK1612/2015). Every patient gave informed consent

## References

1. Savarese G, Becher PM, Lund LH, Seferovic P, Rosano GMC, Coats AJS. Global burden of heart failure: a comprehensive and updated review of epidemiology. Cardiovasc Res. 2023;118(17):3272–3287. doi:10.1093/CVR/CVAC013

2. Mamas MA, Sperrin M, Watson MC, et al. Do patients have worse outcomes in heart failure than in cancer? A primary care-based cohort study with 10-year follow-up in Scotland. Eur J Heart Fail. 2017;19(9):1095–1104. doi:10.1002/EJHF.822

3. Greene SJ, Butler J, Albert NM, et al. Medical Therapy for Heart Failure With Reduced Ejection Fraction: The CHAMP-HF Registry. J Am Coll Cardiol. 2018;72(4):351–366. doi:10.1016/J.JACC.2018.04.070

4. Kapelios CJ, Canepa M, Benson L, et al. Non-cardiology vs. cardiology care of patients with heart failure and reduced ejection fraction is associated with lower use of guideline-based care and higher mortality: Observations from The Swedish Heart Failure Registry. Int J Cardiol. 2021;343:63–72. doi:10.1016/J.IJCARD.2021.09.013

5. Fiuzat M, Ezekowitz J, Alemayehu W, et al. Assessment of Limitations to Optimization of Guideline-Directed Medical Therapy in Heart Failure From the GUIDE-IT Trial: A Secondary Analysis of a Randomized Clinical Trial. JAMA Cardiol. 2020;5(7):1. doi:10.1001/JAMACARDIO.2020.0640

6. Savarese G, Lindberg F, Christodorescu RM, et al. Physician perceptions, attitudes, and strategies towards implementing guideline-directed medical therapy in heart failure with reduced ejection fraction. A survey of the Heart Failure Association of the ESC and the ESC Council for Cardiology Practice. Eur J Heart Fail. 2024;26(6):1408–1418. doi:10.1002/EJHF.3214

7. Ouwerkerk W, Voors AA, Anker SD, et al. Determinants and clinical outcome of uptitration of ACE-inhibitors and beta-blockers in patients with heart failure: a prospective European study. Eur Heart J. 2017;38(24):1883–1890. doi:10.1093/EURHEARTJ/EHX026

8. Peterson PN, Rumsfeld JS, Liang L, et al. Treatment and risk in heart failure: gaps in evidence or quality? Circ Cardiovasc Qual Outcomes. 2010;3(3):309–315. doi:10.1161/CIRCOUTCOMES.109.879478

9. McDonagh TA, Metra M, Adamo M, et al. 2021 ESC Guidelines for the diagnosis and treatment of acute and chronic heart failure. Eur Heart J. 2021;42(36):3599–3726. doi:10.1093/eurheartj/ehab368

10. Cotter G, Deniau B, Davison B, et al. Optimization of Evidence-Based Heart Failure Medications After an Acute Heart Failure Admission: A Secondary Analysis of the STRONG-HF Randomized Clinical Trial. JAMA Cardiol. 2024;9(2):114–124. doi:10.1001/JAMACARDIO.2023.4553

11. F E Harrell Jr, K L Lee, D B Mark. Multivariable prognostic models: issues in developing models, evaluating assumptions and adequacy, and measuring and reducing errors - PubMed. Stat Med. 1996;15(4):361–387. Accessed April 2, 2025. https://pubmed.ncbi.nlm.nih.gov/8668867/

12. Bozkurt B, Coats AJ, Tsutsui H, et al. Universal Definition and Classification of Heart Failure: A Report of the Heart Failure Society of America, Heart Failure Association of the European Society of Cardiology, Japanese Heart Failure Society and Writing Committee of the Universal Definition of Heart Failure. J Card Fail. 2021;27(4):387–413. doi:10.1016/J.CARDFAIL.2021.01.022

13. Crespo-Leiro MG, Metra M, Lund LH, et al. Advanced heart failure: a position statement of the Heart Failure Association of the European Society of Cardiology. Eur J Heart Fail. 2018;20(11):1505–1535. doi:10.1002/EJHF.1236

14. The Consensus Trial Study Group. Effects of Enalapril on Mortality in Severe Congestive Heart Failure. N Engl J Med. 1987;316(23):1429–1435. doi:10.1056/NEJM198706043162301

15. Packer M, Fowler MB, Roecker EB, et al. Effect of carvedilol on the morbidity of patients with severe chronic heart failure: results of the carvedilol prospective randomized cumulative survival (COPERNICUS) study. Circulation. 2002;106(17):2194–2199. doi:10.1161/01.CIR.0000035653.72855.BF

16. Pitt B, Zannad F, Remme WJ, et al. The Effect of Spironolactone on Morbidity and Mortality in Patients with Severe Heart Failure. N Engl J Med. 1999;341(10):709–717. doi:10.1056/NEJM199909023411001/ASSET/33862127-89E0-408B-91B8-654E8D1EF3BA/ASSETS/IMAGES/LARGE/NEJM199909023411001_T4.JPG

17. McMurray JJV, Packer M, Desai AS, et al. Angiotensin–Neprilysin Inhibition versus Enalapril in Heart Failure. N Engl J Med. 2014;371(11):993–1004. doi:10.1056/NEJMoa1409077

18. Brunner-La Rocca HP, Linssen GC, Smeele FJ, et al. Contemporary Drug Treatment of Chronic Heart Failure With Reduced Ejection Fraction: The CHECK-HF Registry. JACC Hear Fail. 2019;7(1):13–21. doi:10.1016/J.JCHF.2018.10.010

19. Ouwerkerk W, Voors AA, Anker SD, et al. Determinants and clinical outcome of uptitration of ACE-inhibitors and beta-blockers in patients with heart failure: a prospective European study. doi:10.1093/eurheartj/ehx113

20. Felker GM, Anstrom KJ, Adams KF, et al. Effect of Natriuretic Peptide–Guided Therapy on Hospitalization or Cardiovascular Mortality in High-Risk Patients With Heart Failure and Reduced Ejection Fraction: A Randomized Clinical Trial. JAMA. 2017;318(8):713–720. doi:10.1001/JAMA.2017.10565

21. Mebazaa A, Davison B, Chioncel O, et al. Safety, tolerability and efficacy of up-titration of guideline-directed medical therapies for acute heart failure (STRONG-HF): a multinational, open-label, randomised, trial. Lancet. 2022;400(10367):1938–1952. doi:10.1016/S0140-6736(22)02076-1

22. Adlbrecht C, Hülsmann M, Wurm R, et al. Outcome of conservative management vs. assist device implantation in patients with advanced refractory heart failure. Eur J Clin Invest. 2016;46(1):34–41. doi:10.1111/ECI.12562

23. Cowie MR, Schöpe J, Wagenpfeil S, et al. Patient factors associated with titration of medical therapy in patients with heart failure with reduced ejection fraction: data from the QUALIFY international registry. ESC Hear Fail. 2021;8(2):861. doi:10.1002/EHF2.13237

24. Greene SJ, Fonarow GC, DeVore AD, et al. Titration of Medical Therapy for Heart Failure With Reduced Ejection Fraction. J Am Coll Cardiol. 2019;73(19):2365–2383. doi:10.1016/J.JACC.2019.02.015

25. Harrington J, Fonarow GC, Khan MS, et al. Medication-Attributable Adverse Events in Heart Failure Trials. JACC Hear Fail. 2023;11(4):425–436. doi:10.1016/J.JCHF.2022.11.026

26. Grobbee DE, Filippatos G, Desai NR, et al. Epidemiology and risk factors for hyperkalaemia in heart failure. ESC Hear Fail. 2024;11(4):1821. doi:10.1002/EHF2.14661

27. Butler J, Anker SD, Lund LH, et al. Patiromer for the management of hyperkalemia in heart failure with reducedejection fraction: the DIAMOND trial. Eur Heart J. 2022;43(41):4362. doi:10.1093/EURHEARTJ/EHAC401

28. Weir MR, Rossignol P, Pitt B, et al. Patiromer-Facilitated Renin-Angiotensin-Aldosterone System Inhibitor Utilization in Patients With Heart Failure With or Without Comorbid Chronic Kidney Disease: Subgroup Analysis of DIAMOND Randomized Trial. Am J Nephrol. Published online August 19, 2024. doi:10.1159/000540453

29. Bhandari S, Mehta S, Khwaja A, et al. Renin–Angiotensin System Inhibition in Advanced Chronic Kidney Disease. N Engl J Med. Published online December 1, 2022. doi:10.1056/NEJMOA2210639

30. Guidetti F, Lund LH, Benson L, et al. Safety of continuing mineralocorticoid receptor antagonist treatment in patients with heart failure with reduced ejection fraction and severe kidney disease: Data from Swedish Heart Failure Registry. Eur J Heart Fail. 2023;25(12):2164–2173. doi:10.1002/EJHF.3049

31. The EMPA-KIDNEY Collaborative Group, Herrington WG, Staplin N, et al. Empagliflozin in Patients with Chronic Kidney Disease. N Engl J Med. 2023;388(2):117–127. doi:10.1056/NEJMoa2204233

32. Damman K, Gori M, Claggett B, et al. Renal Effects and Associated Outcomes During Angiotensin-Neprilysin Inhibition in Heart Failure. JACC Hear Fail. 2018;6(6):489–498. doi:10.1016/J.JCHF.2018.02.004

33. Le D, Grams ME, Coresh J, Shin JI. Sacubitril-Valsartan in Patients Requiring Hemodialysis. JAMA Netw Open. 2024;7(8):e2429237–e2429237. doi:10.1001/JAMANETWORKOPEN.2024.29237

